# Deep phenotyping obesity using EHR data: Promise, Challenges, and Future Directions

**DOI:** 10.1101/2024.12.06.24318608

**Authors:** Xiaoyang Ruan, Shuyu Lu, Liwei Wang, Andrew Wen, Murali Sameer, Hongfang Liu

## Abstract

Obesity affects approximately 34% of adults and 15–20% of children and adolescents in the U.S, and poses significant economic and psychosocial burdens. Due to the multifaceted nature of obesity, currently patient responses to any single anti-obesity medication (AOM) vary significantly, highlighting the need for developing approaches to obesity deep phenotyping and associated precision medicine. While recent advancement in classical phenotyping-guided pharmacotherapies have shown clinical value, they are less embraced by healthcare providers within the precision medicine framework, primarily due to their operational complexity and lack of granularity. From this perspective, several recent review articles highlighted the importance of obesity deep phenotyping for personalized precision medicine. In view of the established role of electronic health record (EHR) as an important data source for clinical phenotypings, we offer an in-depth analysis of the commonly available data elements from obesity patients prior to pharmacotherapy. We also experimented with a multi-modal longitudinal deep autoencoder to explore the feasibility, data requirements, clustering patterns, and challenges associated with EHR-based obesity deep phenotyping. Our analysis indicates at least nine clusters, among which five have distinct explainable clinical relevance. Further research within larger independent cohorts to validate the reproducibility, uncover more detailed substructures and corresponding treatment response is warranted.

**Background:** Obesity affects approximately 40% of adults and 15–20% of children and adolescents in the U.S, and poses significant economic and psychosocial burdens. Currently, patient responses to any single anti-obesity medication (AOM) vary significantly, making obesity deep phenotyping and associated precision medicine important targets of investigation.

**Objective:** To evaluate the potential of EHR as a primary data source for obesity deep phenotyping, we conduct an in-depth analysis of the data elements and quality available from obesity patients prior to pharmacotherapy, and apply a multi-modal longitudinal deep autoencoder to investigate the feasibility, data requirements, clustering patterns, and challenges associated with EHR-based obesity deep phenotyping.

**Methods:** We analyzed 53,688 pre-AOM periods from 32,969 patients with obesity or overweight who underwent medium- to long-term AOM treatment. A total of 92 lab and vital measurements, along with 79 ICD-derived clinical classifications software (CCS) codes recorded within one year prior to AOM treatment, were used to train a gated recurrent unit with decay based longitudinal autoencoder (GRU-D-AE) to generate dense embeddings for each pre-AOM record. principal component analysis (PCA) and gaussian mixture modeling (GMM) were applied to identify clusters.

**Results:** Our analysis identified at least nine clusters, with five exhibiting distinct and explainable clinical relevance. Certain clusters show characteristics overlapping with phenotypes from traditional phenotyping strategy. Results from multiple training folds demonstrated stable clustering patterns in two-dimensional space and reproducible clinical significance. However, challenges persist regarding the stability of missing data imputation across folds, maintaining consistency in input features, and effectively visualizing complex diseases in low-dimensional spaces

**Conclusion:** In this proof-of-concept study, we demonstrated longitudinal EHR as a valuable resource for deep phenotyping the pre-AOM period at per patient visit level. Our analysis revealed the presence of clusters with distinct clinical significance, which could have implications in AOM treatment options. Further research using larger, independent cohorts is necessary to validate the reproducibility and clinical relevance of these clusters, uncover more detailed substructures and corresponding AOM treatment responses.

## Background

Obesity affects ∼40% of adults and ∼15–20% of children and adolescents in the U.S. ^1^. It is projected that over 85% of adults will be overweight or obese by 2030 in the USA ^2^. Obesity increases the risk of a wide spectrum of chronic diseases and causes profound economic and psychosocial burden. Obesity arises from a complex interplay of genetic ^3,4,5,6^, non-genetic ^7,8,9^, and epigenetic factors ^10,11,12,13^. Due to this multifaceted nature, no single therapy, either non-invasive (e.g. anti-obesity medications (AOMs), dietary control, hydrogel) or invasive (e.g. bariatric surgeries), can robustly predict patient response. For example, meta-analysis indicates wide interindividual response after various bariatric surgeries during longer-term follow-up ^14,15^. Roux-en-Y gastric bypass of 6000 patients showed 23.1% as non-responders 5 years after surgery ^16^. Follow-up of 652 patients with sleeve gastrectomy for 7 years indicated 27.8% weight recidivism ^17^. For AOMs, even the most effective GLP1 drug Tirzepatide claimed 10% weight loss in only 69% participants after 1 year at a relatively safe dosage ^18^. Other AOMs ^19^ and treatment strategies like hydrogels ^20^ and vagal nerve blockade ^21,22^ generally show broader interindividual variations, which make them often an auxiliary part of weight management plan ^23–25^.

The inconsistent therapeutic response makes obesity phenotyping (i.e. classify obesity into subtypes) and associated precision management important targets of investigation. Earlier advancements in obesity staging ^26^ and phenotyping-guided pragmatic trial ^27^ have moved beyond the oversimplified classification by Body Mass Index (BMI) ^28^ and demonstrated clinical values ^29,30,31^. However, healthcare practitioners (HCPs) still found major obstacles in adopting them for obesity precision medicine ^32^. One obstacle lies in the complexity of these strategies. For example, the trial by ^27^ involves tedious (e.g. satiation measured by ad libitum buffet meal) and subjective (e.g. hunger measured by visual analog scale) measurement processes. There are also considerable gaps between the desired and actual level of acceptance of obesity treatment guidelines by HCPs ^42,33,34^. Another major obstacle is the lack of granularity of the phenotypic information to personalized context ^32^. Phenotype-based pharmacotherapy has been successful only in rare monogenic obesity ^35^. To date, the sole predictor of the long-term response of pharmacotherapy is the short-term weight loss result, which is useless for guiding the initial treatment selection ^36^.

Recent review articles ^37,38^ concluded with the necessity of obesity deep phenotyping for precision medicine, as “when considering obesity, every person should be assessed based on their own specific and unique circumstances” ^32^. Echoing this statement, our recent study covering more than 1 million obesity/overweight patients indicates wide inter individual responses to any single AOM regardless of exposure lengths ^39^. To date, there is a significant dearth of research on important questions like in what clinical context (e.g. at the time of diagnosis; before initiating pharmacotherapy; postsurgical weight management) should we discuss “deep phenotyping”, the type/source/availability/quality of relevant data elements, the computational methodologies, and how “deep” should and can we go. To take a peek into this intricate and multifaceted research question, here we narrow down the scope to deep phenotyping before initiating pharmacotherapy, a period with direct relation to drug response and thus potential clinical impact. We propose that electronic health records (EHRs) are a valuable data source for this purpose, as they provide detailed information across patient journeys and have inspired relevant research ^40,41^. Using real-world EHR data from 444,219 patients with obesity or overweight diagnosed between 2005 and 2023, we analyzed commonly available data elements and their quality before pharmacotherapy. We also tested a multi-modal longitudinal deep autoencoder to examine the feasibility, data requirements, clustering patterns, and challenges of EHR-based obesity deep phenotyping.

## Materials and Methods

### EHR database and the case cohort

The study is based on EHR data from the outpatient practice of the University of Texas Health Sciences Center at Houston’s McGovern Medical School. The data is transformed (from Allscripts Touchworks EHR pre-2021, GE Centricity EHR pre-2021 for billing, EPIC EHR post-2021) to OMOP common data model (CDM) on a nightly basis to harmonize data query format. Currently the OMOP CDM instance covers 6.5M patients, among which ∼3.7M patients have condition(s) documented between 2005/01/01 and 2023/12/31. From these patients we identified 456,890 patients with either obesity (n=217,040) (Supp Text 1) or overweight (n=239,850) diagnosis. Overweight is defined as patients with certain comorbidities (Supp Text 2) within 1 year prior to a BMI measurement ≥27. From the 456,890 patients we defined the case cohort as the 32,969 patients with medium (> 112 days) to long exposure to AOMs and have no bariatric surgeries. The University of Texas Institutional Review Board (IRB) approved the study, and the Ethics Committee waived the need for written informed consent from participants.

### FDA approved AOMs (F-AOMs) and Off-Label AOMs (O-AOMs)

For F-AOMs, we considered bupropion naltrexone, orlistat, phentermine, phentermine topiramate, liraglutide, semaglutide, tirzepatide that were approved by FDA for obesity treatment. For O-AOMs we considered bupropion, canagliflozin, dapagliflozin w/wo saxagliptin, empagliflozin w/wo linagliptin, lisdexamfetamine, metformin w/wo (liflozin, liptin, litazone, statin), topiramate, zonisamide. Specifically, exposure to an O-AOM ingredient for less than 30 days without a neighboring (30 days before or after) record was removed to allow for occasional use of O-AOMs for other purposes. While most of the O-AOMs are for diabetes treatment, we make no explicit distinction between the treatment purposes when the treatment course is longer than 30 days, in which case an impact on body weight should be anticipated. For each drug ingredient, we searched the RxNav ^42^ database for all aliases. We then searched the Observational Health Data Sciences and Informatics (OHDSI) Athena ^43^ database for concept ids of the original ingredient and all aliases. The concept ids were combined to represent a single corresponding ingredient.

### Definition of AOM treatment session

An AOM treatment session represents a continuous period of exposure to generally the same active ingredient(s). Specifically, compare to an existing exposure record *REC_A_*, a new exposure record *REC_B_* belongs to the same treatment session (as *REC_A_*) if it 1) has exactly the same ingredient(s) as *REC_A_* and there is ≤40 days gap between start of *REC_B_* and end of *REC_A_*, or 2) has less ingredient than *REC_A_*, and has both exposure length and gap to *REC_A_* ≤40 days, or 3) has new ingredient(s) added within 40 days of initiating *REC_A_* and has exposure length ≤40 days. For all other circumstances, we consider *REC_B_* as a new treatment session. For the 32,969 patients in the case cohort, we identified a total of 53,688 AOM treatment sessions.

### Pre-AOM period, and data points sampling scheme

We defined “pre-AOM period” as 1 year (i.e. 365 days) before starting each AOM treatment session. Thus, there are 53,688 pre-AOM periods corresponding to each of the 53,688 AOM treatment sessions. For each pre-AOM period, we sampled data points every 30 days with a maximum look-back period of 30 days, resulting in 13 sampling points and 12 intervals for each feature. Notably, the interval closest to treatment session start date spans from 35 days to 5 days before initiation. Throughout the rest of the text, we refer to the date 4 days before AOM initiation as the index date. We designed a 4-day lead time to allow for any final data collection or assessments that can be completed without interfering with the treatment process.

### Normal BMI control period

We randomly selected 10,000 case pre-AOM periods to match with corresponding normal BMI control periods, of patients with no obesity or overweight diagnosis and no BMI >25 kg/m². The matching process followed these criteria: 1) the case and control periods were matched by gender, 2) the age at the normal BMI measurement was within one year of the case’s age at AOM initiation, and 3) the normal BMI measurement date was within one year of the case’s AOM initiation date. In the end, 8,773 case pre-AOM periods were successfully matched one-to-one with normal BMI control periods that belonged to 8,185 normal BMI patients.

### Explored features

#### Measurements

We examined all numerical measurements recorded in the OMOP “measurement” table, which includes all lab and vital records (referred as “measurements” in the remaining text) from the original EHR databases. The units of each measurement were harmonized to the primary unit type through manual inspection. Specifically, data quality of each pre-AOM period was defined as the average proportion of observed measurements of the 13 sampling points before AOM initiation.

#### Diagnosis code

All ICD-9 diagnosis codes were cast into 265 categories of clinical classifications software (CCS) codes ^44^. All ICD-10 diagnosis codes were mapped to ICD-9 code through general equivalence mapping provided by Center for Medicare & Medicaid Services. CCS is a tool for clustering patient ICD-9 diagnoses and procedures into a manageable number of clinically meaningful categories for easy presentation and statistical analysis. The CCS codes were one hot encoded before presenting to the autoencoder model.

### Embedding of EHR data

#### Embedding of longitudinal EHR data

We re-engineered the GRU-D ^45^ based longitudinal deep learning architecture to function as an autoencoder (GRU-D-AE) for embedding the longitudinal EHR data from pre-AOM period. The architecture is illustrated in Figure 1a, where *X_t_* represents all input data at timestep *t*, *x_t_* is the normalized feature value (e.g., converting lab measurements to corresponding z-scores; scaling age to a 0-1 range), *m_t_* is the missing indicator (0 for missing and 1 for presence), and *d_t_* is the time since the last actual observation. The bottleneck layer *h_T_* contains the dense embedding vector as the output of GRU-D from the last time step *T*. *h_T_* is then passed to a native RNN network (in this case, a native GRU model) to generate the reconstructed feature value *x̅*. Figure 1b shows the data processing flow, from raw EHR data to principal components, and how GRU-D-AE functions in the process.

**Figure 1.**
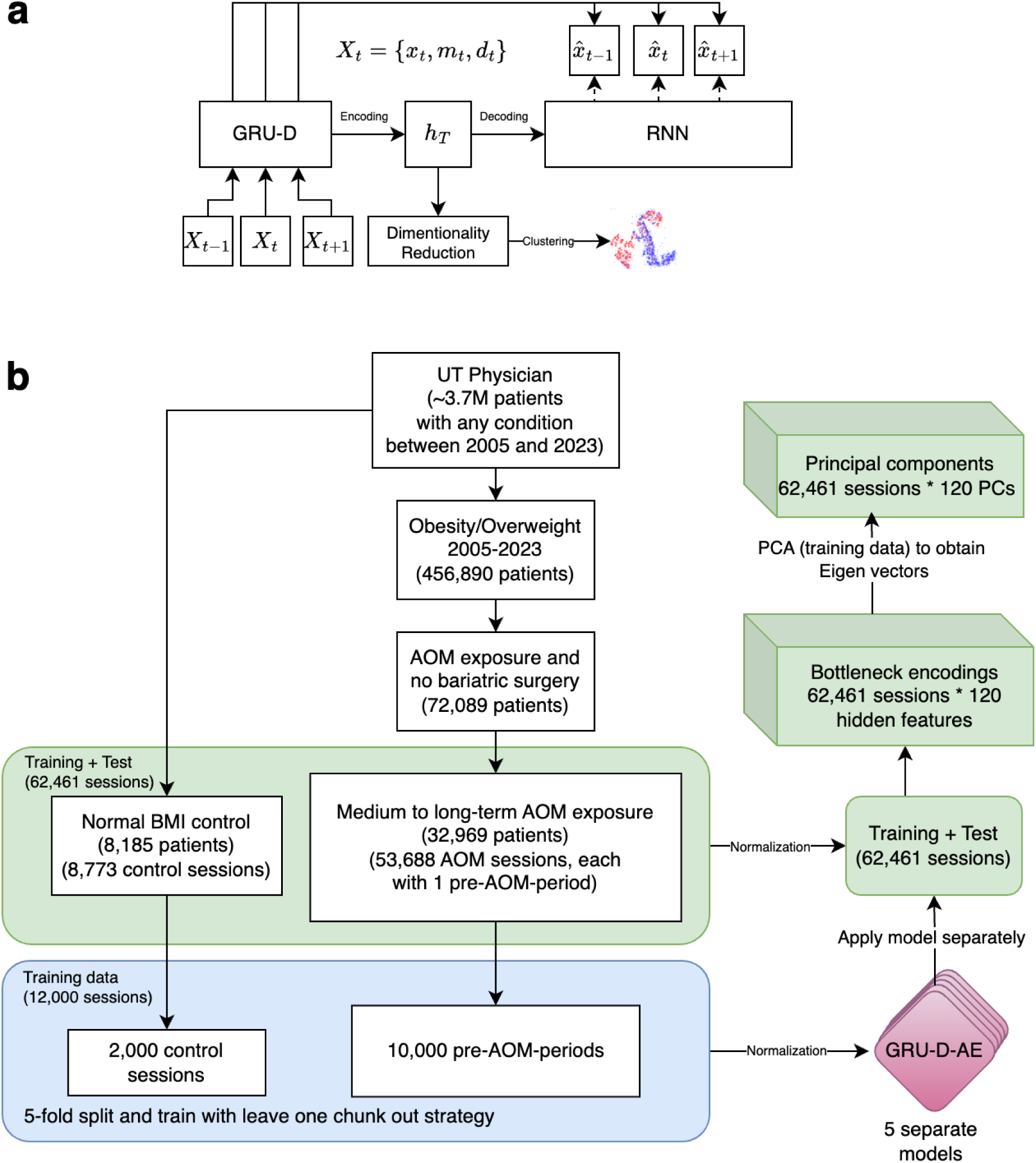
a) GRU-D-AE architecture. b) Sample processing flow and the generation of principal components (PCs) from longitudinal EHR.

Specifically, the loss function for GRU-D-AE is expressed as

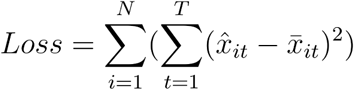

Where *N* represents the number of training samples, *T* the longitudinal time steps during pre-AOM period, *x̅_it_* the reconstructed feature value for sample *i* at time step *t*, *x̂_it_* the original observation or, if missing, an imputed value through the GRU-D missing parameterization mechanism.

#### Embedding of static-transformed longitudinal EHR data

As a baseline comparison, we conducted experiments using an atemporal sparse autoencoder (SAE) to embed static-transformed longitudinal EHR data. The static transformation involved extracting the last observed feature within the 1-year period preceding AOM initiation. Features with no observations during this period were imputed with 0 and excluded from the loss function through masking. The experiment pipeline, including the training sample indices, was identical to that used in the GRU-D-AE model (Fig. 1b).

### Principal component analysis (PCA) and clustering algorithm

PCA was conducted on the embeddings generated from the training data to derive eigenvectors. These eigenvectors were then applied to the embeddings from both the training and test datasets to calculate the corresponding principal components (PCs) (Figure 1b). We used the Gaussian Mixture Model (GMM) to perform probability-based clustering on the top 40 PCs of each embedding. To visualize the embeddings in two-dimensional space, we explored two methods: a scatterplot of the top two PCs and a t-SNE plot of the top 40 PCs.

### Computational environment

The computational environment for data analysis consisted of a Unix-based high-performance computing system with 192 CPUs. Deep learning models were developed using PyTorch version 2.3.1, implemented in Python version 3.12.2. Statistical analyses were conducted using R version 4.3.3. PCA was performed with the built-in *stats* package in R, while GMM was implemented using the *mclust* package (version 6.1.1). t-SNE dimensionality reduction was carried out with the *Rtsne* package (version 0.17). Data visualization tasks were completed using the *ggplot2* package (version 3.5.0) in R.

## Results

### Baseline characteristics of the study population

From 2005/01/01 to 2023/12/31, we identified 444,309 (12%) patients with either obesity (n=204,688) or overweight (n=239,621) diagnosis. Among these patients 72,089 (16%) were exposed to AOM therapy, with a total of 136,728 AOM treatment sessions (i.e. averagely ∼2 treatment sessions for each patient). 53,688 (39%) of the AOM sessions from 32,969 patients belong to medium or long term exposure (>=112 days). The baseline characteristics of the study population were shown in Table 1. The sample processing flow is illustrated in Figure 1b.

**Table 1.**
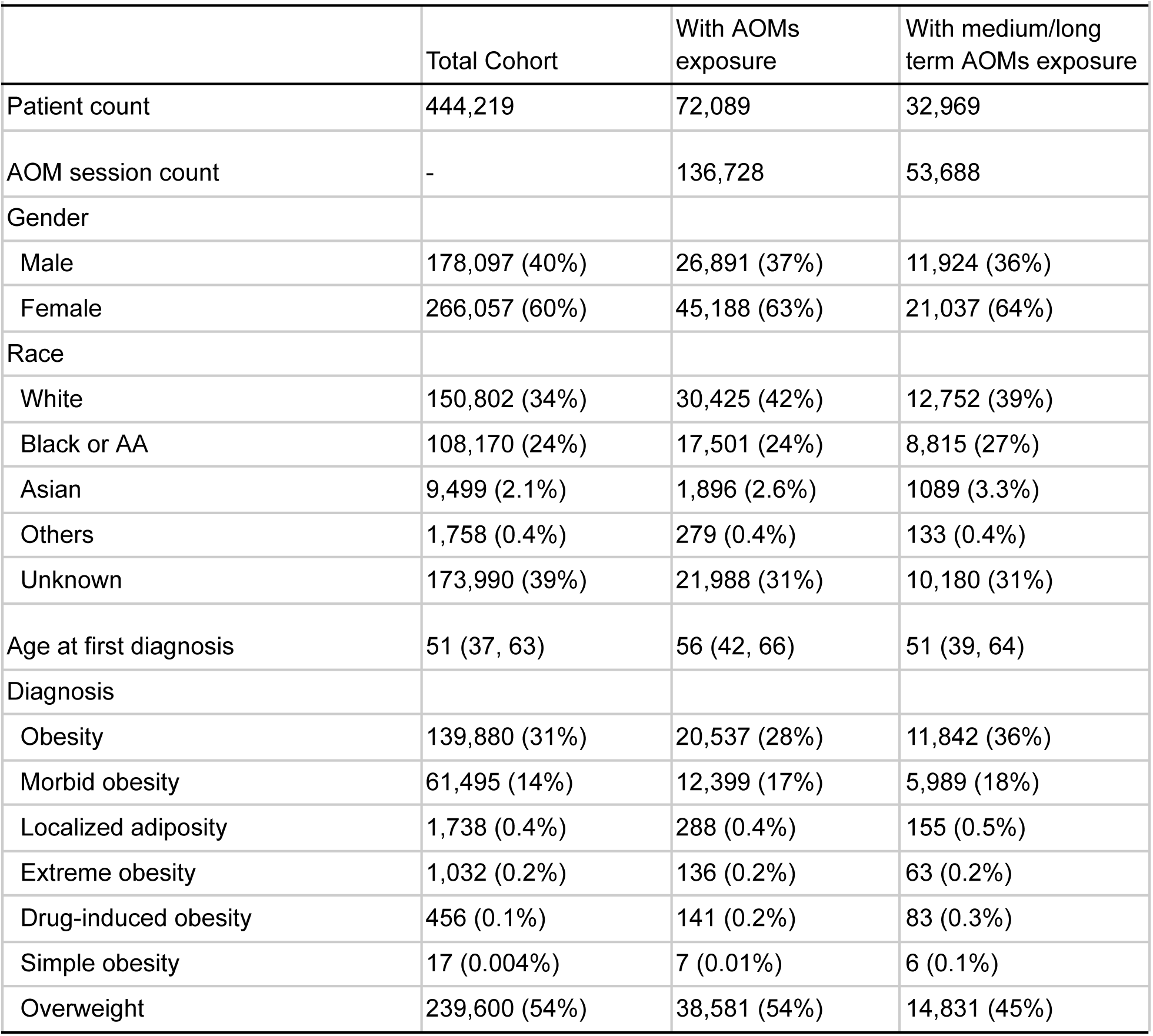
Demographics of the study population.

|  | Total Cohort | With AOMs exposure | With medium/long term AOMs exposure |
| --- | --- | --- | --- |
| Patient count | 444,219 | 72,089 | 32,969 |
| AOM session count | - | 136,728 | 53,688 |
| Gender |  |  |  |
| Male | 178,097 (40%) | 26,891 (37%) | 11,924 (36%) |
| Female | 266,057 (60%) | 45,188 (63%) | 21,037 (64%) |
| Race |  |  |  |
| White | 150,802 (34%) | 30,425 (42%) | 12,752 (39%) |
| Black or AA | 108,170 (24%) | 17,501 (24%) | 8,815 (27%) |
| Asian | 9,499 (2.1%) | 1,896 (2.6%) | 1089 (3.3%) |
| Others | 1,758 (0.4%) | 279 (0.4%) | 133 (0.4%) |
| Unknown | 173,990 (39%) | 21,988 (31%) | 10,180 (31%) |
| Age at first diagnosis | 51 (37, 63) | 56 (42, 66) | 51 (39, 64) |
| Diagnosis |  |  |  |
| Obesity | 139,880 (31%) | 20,537 (28%) | 11,842 (36%) |
| Morbid obesity | 61,495 (14%) | 12,399 (17%) | 5,989 (18%) |
| Localized adiposity | 1,738 (0.4%) | 288 (0.4%) | 155 (0.5%) |
| Extreme obesity | 1,032 (0.2%) | 136 (0.2%) | 63 (0.2%) |
| Drug-induced obesity | 456 (0.1%) | 141 (0.2%) | 83 (0.3%) |
| Simple obesity | 17 (0.004%) | 7 (0.01%) | 6 (0.1%) |
| Overweight | 239,600 (54%) | 38,581 (54%) | 14,831 (45%) |

### EHR feature availability

#### Overall feature availability during pre-AOM period

During the pre-AOM period, which has potentially the most relevant data for deep phenotyping, we identified 92 measurements with >=5% overall presence rate and non-zero standard deviation (Supp Table 1). The measurements with >=70% presence rate are body weight, body surface area (BSA), BMI, height, blood pressure (BP), and heart rate (HR). Whereas lab measurements for basic metabolic panels (e.g. glucose, calcium, sodium), and lipid levels (e.g. HDL-C, LDL-C, triglyceride) generally have around or >=50% presence rate. Examining the distribution of overall feature presence rates among patient subgroups reveals no differences between males and females, nor across racial groups (i.e., White, Black or African American, Asian, etc).

Out of 285 CCS categories, we identified 79 with >=5% overall presence rate (Supp Table 2). The most common CCS categories are diabetes mellitus (DM) without complications (63%), hypertension (55%), hypertension with complications (55%), and hyperlipidemia (51%). Nutritional disorders, including any type of obesity diagnosis, were present in 40% of cases within one year prior to the AOM session, generally aligning with the percentage (46%) of obesity patients in our case cohort (i.e. obesity + overweight).

#### Temporal windows based feature presence rate

We sampled features every 30 days within one year surrounding the treatment session initiation date to examine the feature presence rates across the 24 corresponding time intervals. For both measurements (Figure 2, Supp Figure 1) and CCS codes (Figure 2, Supp Figure 2), the feature presence rates were remarkably higher in the time interval immediately before the index date (i.e., Day −35 to −5), and slightly elevated around 180, 90, and 30 days before the index date. This pattern of increased feature presence was not evident after the index date, and was not seen in the normal BMI control periods (data not shown). For the pre-AOM periods, 50% measurements were contributed by 22% treatment sessions. On the other hand, 9.9% patients and 11% treatment sessions have no single measurement during this period.

**Figure 2.**
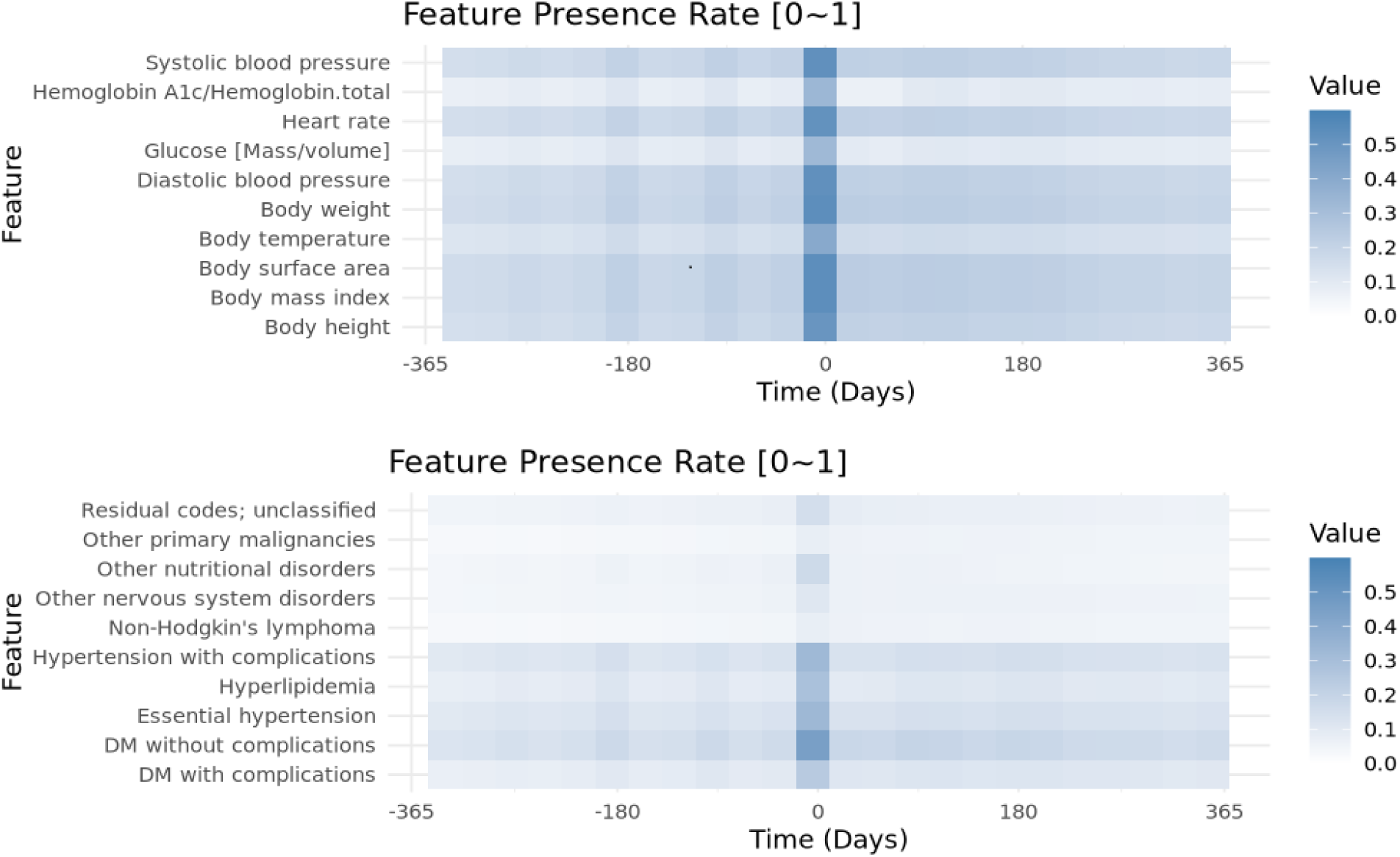
Feature presence rate of the top 10 a) Measurements b) CCS codes appeared within 1 year before and after initiating medium to long term AOM therapy.

### Feature presence rate throughout years

Analyzing feature presence rates from 2005 to 2023 revealed a general upward trend for both measurements and CCS codes over the years. Specifically, the Mann-Kendall non-parametric trend test showed that all 92 measurements have significantly increased presence rates (p ≤ 0.05) over time. The top five measurements with the highest increase are microalbumin, creatinine in urine, HDL-C, total cholesterol, and ferritin in serum or plasma. Additionally, 50 out of the 79 CCS categories demonstrated a significant increase in presence rates over the years, with the top five being anxiety disorders, nutritional deficiencies, other acquired deformities, other non-traumatic joint disorders, and screening and history of mental health and substance abuse codes (Supp Figure 3). Two CCS categories—chest pain and skin and subcutaneous tissue infections—showed a significant (p ≤ 0.05) or marginally significant (p ≤ 0.1) decrease in presence rates over the years.

### Encoding and reconstruction of longitudinal EHR data

We experimented with various bottleneck layer neuron sizes (n=40, 60, 120) for the GRU-D-AE model. Generally, larger bottleneck sizes allow the autoencoder to capture more intricate temporal structures. Supp Figure 4 shows the GRU-D-AE based reconstructions of 171 temporal features (92 measurements and 79 CCS codes) from a single patient, using 5-fold cross-validation with a bottleneck layer of 120 neurons. Comparing results across the 5 models indicates slightly different imputation patterns across temporal steps and successful reconstruction of input feature patterns at high levels. For the remainder of the paper, we present results exclusively based on 120 bottleneck layer size, as this configuration demonstrated optimal performance.

### Clustering of case pre-AOM periods

By encoding the pre-AOM periods and projecting them onto principal component (PC) spaces, we visualized clustering patterns in lower-dimensional spaces, as shown in Figure 3. Specifically, Figure 3a highlights the distribution of pre-AOM periods across different data quality quartiles. Data points from the bottom quartile (red) are densely packed in the lower-left corner, while those from the top quartile (purple) are more dispersed and form distinct clusters. The T-SNE plot with top 40 PCs (explained 80% variance) reveals a complete separation of bottom quartile data points and partial separation of 2nd quartile data points. Importantly, applying the same pipeline (Figure 1b) to each of the 5-fold models shows highly reproducible clustering patterns, with different projection angles (Supp Figure 5). After removing low-quality pre-AOM periods (i.e., the 1st and 2nd quartiles), the remaining high-quality periods (i.e., the 3rd and 4th quartiles) form clusters that are less relevant to data quality (Figure 3b). GMM clustering of the 40 PCs of the high-quality periods suggests at least 9 distinct clusters within the case group (Figure 3c). Notably, GMM-based clusters may align (e.g., clusters 2, 5, 7) or differ (e.g., clusters 1, 6, 9) from the visually identifiable cluster centers based on the two PCs (Supp Figure 6). The visual separation between clusters improves slightly with T-SNE-based dimensionality reduction (Supp Figure 7). When the clustering results are colored by traditional obesity diagnosis categories (e.g., drug-induced obesity, extreme obesity, localized adiposity, morbid obesity), no clear relationship emerges between traditional obesity diagnoses and the EHR-based clustering of pre-AOM periods (Supp Figure 8).

**Figure 3.**
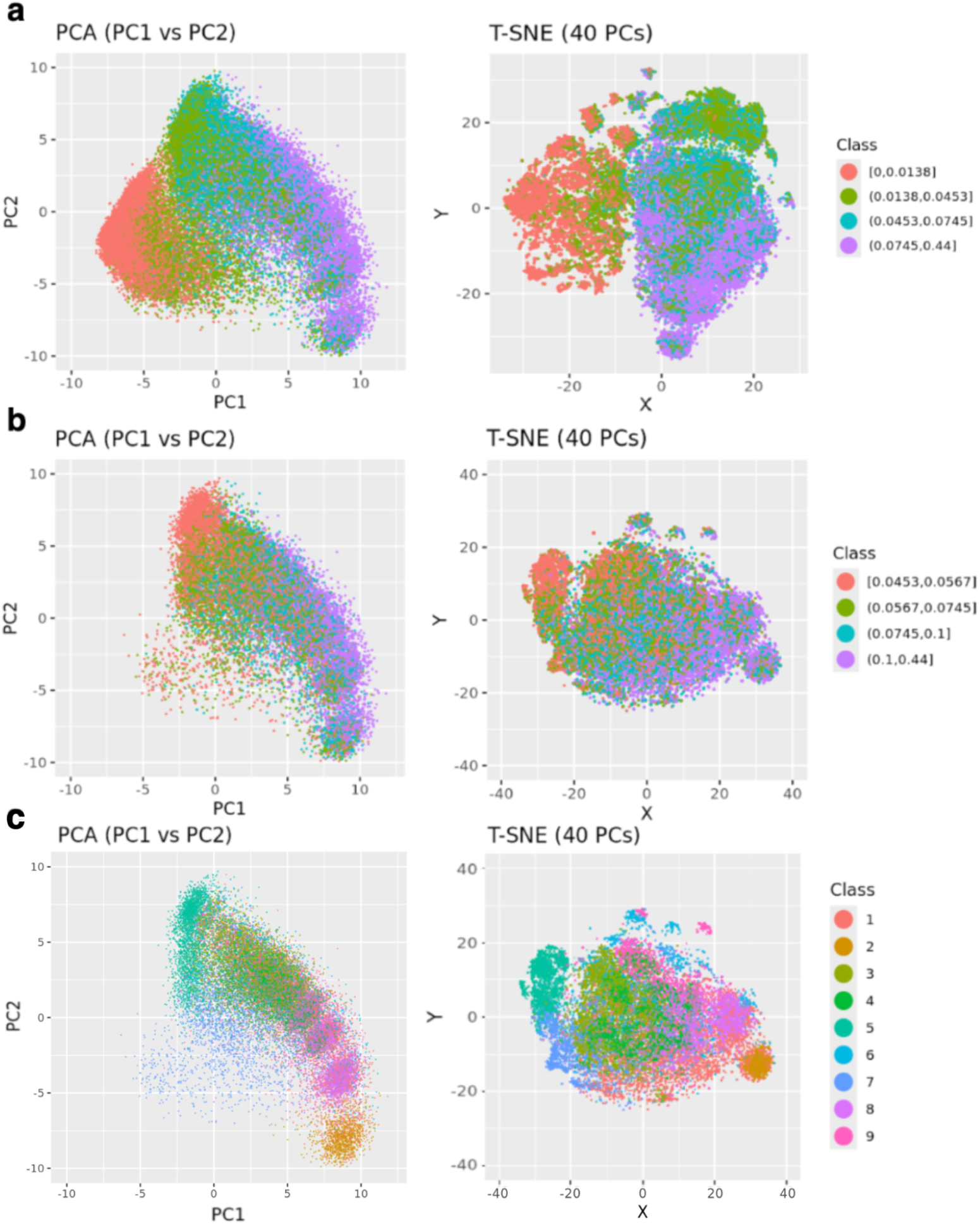
GRU-D-AE-based clustering of case pre-AOM periods. a) Coloring all pre-AOM periods according to data quality quartiles. b) Low-quality data points (below the median) are removed, with the remaining points colored by data quality quartiles. c) Colored by GMM-based clustering of the high quality data points (PCA plot was dimmed to highlight cluster center).

### Clustering of case versus control periods

Visualization of the top two PCs and T-SNE plot (Figure 4) from the case pre-AOM periods and the control periods showed clear separation of the majority of control periods from the high-quality case periods. However, a minor proportion of the control periods formed clusters with centers overlapping the case group on the top two PCs (Figure 4a), with no clear pattern of overlap on the T-SNE plot (Figure 4b).

**Figure 4.**
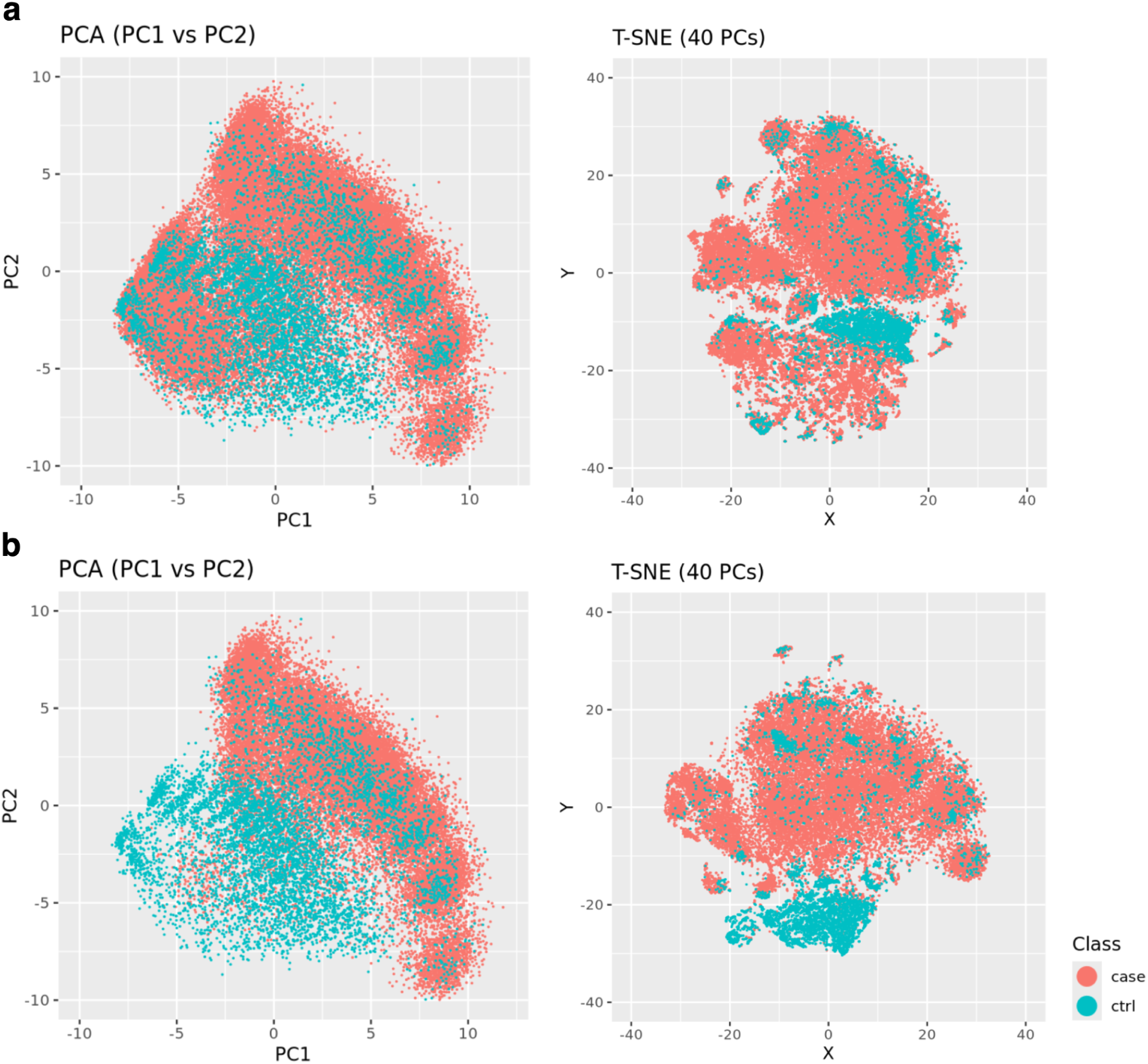
Clusters of case pre-AOM periods versus controls. a) All case data points displayed. b) Remove low quality data points from the case group.

### Clustering of multiple pre-AOM periods from the same patient

For patients with multiple pre-AOM periods from corresponding multiple AOM treatment sessions, their clustering behavior is influenced by both data quality and intrinsic physiological factors reflected through measurements and diagnosis status. In Figure 5, all three patients exhibit improved data quality over time (later pre-AOM periods are indicated by larger dots, which generally have better data quality), with varying degrees of clustering tightness in the high-quality pre-AOM periods (represented by cyan and purple points).

**Figure 5.**
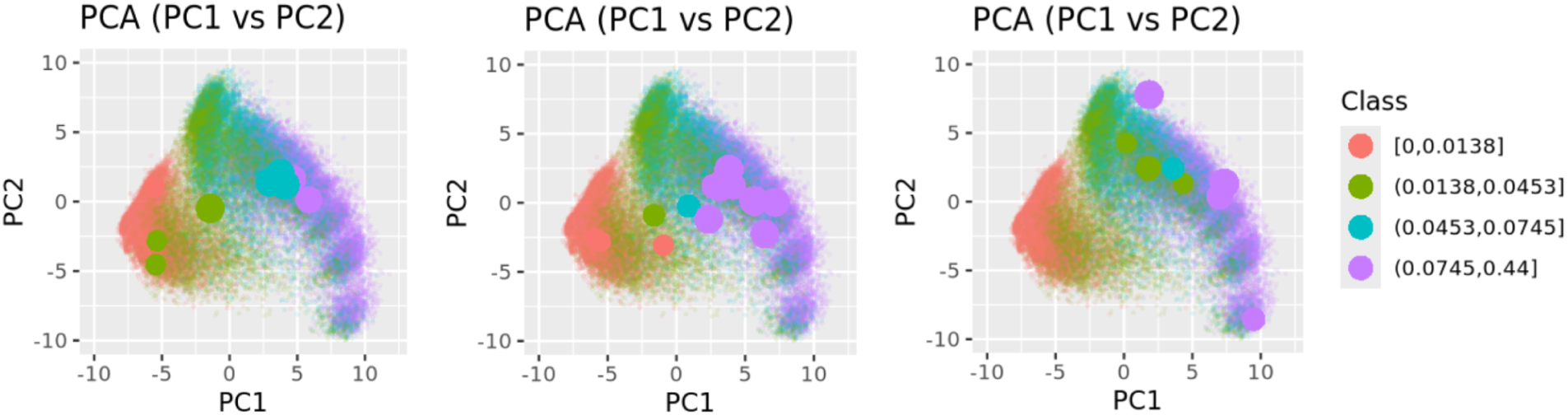
Illustration of three patients, each with multiple AOM sessions and respective pre-AOM periods. Smaller point sizes indicate pre-AOM periods earlier in time, while colors represent different data quality quartiles.

### Characterizing GMM-based clusters of pre-AOM periods

We conducted two-way clustering of the GMM-based clusters of pre-AOM periods against CCS prevalence rates (Figure 6), average measurement values (Figure 7), and temporal measurement presence rate (Supp Figure 9). Here we briefly describe the 5 clusters with remarkable clinical relevance.

**Figure 6.**
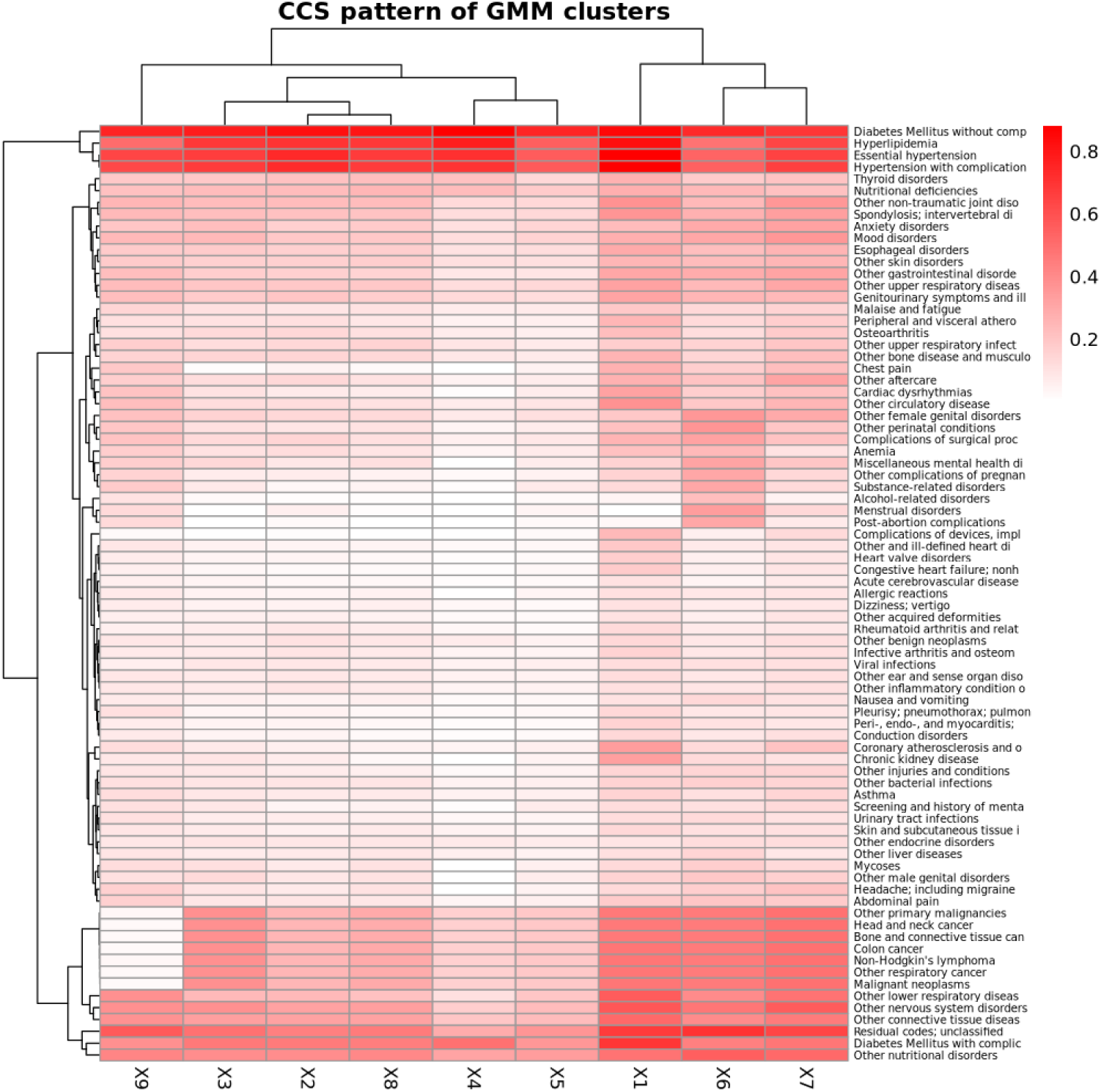
Two-way clustering of CCS diagnosis prevalence rates against GMM-based clusters of pre-AOM periods. Prevalence rates are calculated based on any diagnoses made within one year prior to the index date.

**Figure 7.**
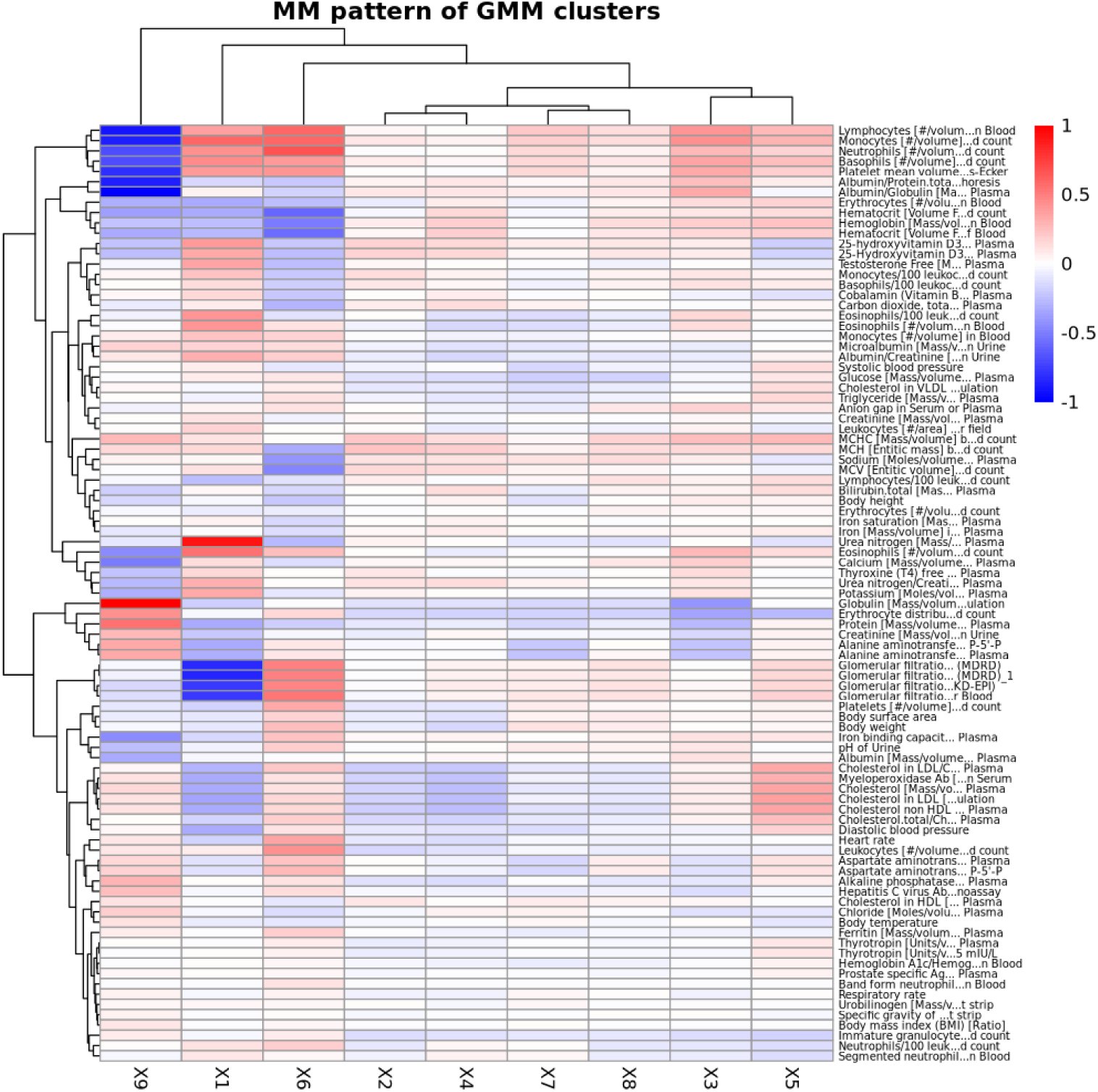
Two-way clustering of mean measurement values (z-score transformed) against GMM-based clusters of pre-AOM periods. The measurement values represent the most recent observations within one year prior to the index date.

The **1st cluster** is distinguished by the highest prevalence of chronic kidney disease (CKD), coronary atherosclerosis, and diabetes mellitus with complications. This cluster also shows notably low glomerular filtration rates, elevated blood urea nitrogen, and comparatively lower levels of low-density cholesterols and myeloperoxidase.

The **5th cluster** is primarily characterized by the highest levels of low-density lipoprotein (LDL), very low-density lipoprotein (VLDL), triglyceride, systolic/diastolic blood pressure (SBP, DBP), and myeloperoxidase among all clusters. This cluster also has slightly higher glomerular filtration rates than other clusters except cluster 6. However, its disease prevalence rates are not significantly distinct from those of other clusters.

The **6th cluster** is marked by a higher prevalence of reproductive and genital health disorders (e.g., female genital disorders, menstrual disorders, and post-abortion complications). It also features higher heart rates, the highest body weight and glomerular filtration rates, and increased levels of leukocytes, lymphocytes, monocytes, neutrophils, and glucose levels.

The **7th Cluster** is characterized by a high level of primary malignancies (e.g. head and neck cancer, bone and connective tissue cancer, and colon cancer), the highest presence rate of temporal anthropometric and physiological measurements (e.g. body height, body weight, body temperature, BP, heart rate), and a lower to average presence rate of other more advanced lab tests. It is also featured with a mildly higher body weight.

The **9th cluster** is notable for significantly lower prevalence rates of primary malignancies, and relatively lower leukocyte levels (including lymphocytes, monocytes, and neutrophils) alongside elevated globulin levels.

No notable difference was observed between the clusters on average ADI-based SDOH factors including median household income, mean education percentage, and mean insurance percentage (data not shown). Inspecting demographic characteristics (i.e. age, gender, race) indicates a relatively lower male proportion in cluster 6, slightly higher proportion of black people in cluster 9, and slightly older age in cluster 1 (data not shown).

Interestingly, clusters characterized by distinct clinical traits (e.g., reproductive and genital health disorders in Cluster 6 and the absence of primary cancer in Cluster 9) were not visually distinguishable on the first two PCs (Supp Figure 6) and only formed vague boundaries in the 40-PC based T-SNE plot (Supp Figure 7). In contrast, clusters that showed clear separation in the low-dimensional spaces (e.g., Cluster 2) may not have distinct clinical characteristics as other clusters, at least based on the available study features. We also tried GMM clustering using just the top two PCs, which produced visually distinguishable clusters but lacked meaningful clinical differentiation (data not shown).

### SAE-based embedding and clustering of pre-AOM periods

As a baseline comparison, we evaluated the SAE-based embedding of static-transformed pre-AOM periods and summarize the results here. Using the same bottleneck layer size (n=120), the SAE model reliably reconstructed the static-transformed features (Supp Figure 11). However, embeddings generated by the SAE model do not exhibit visually distinct clusters on the top two PCs (Supp Figure 12a). When colored by data quality quartiles, pre-AOM periods with lower data quality appear less separable (Supp Figure 12a), aligning with the results observed in the GRU-D-AE model (Figure 4a). Additionally, SAE-based embeddings show weaker differentiation between pre-AOM periods across different data quality quartiles (Supp Figure 12). For high-quality pre-AOM periods, GMM clustering reveals less defined cluster boundaries on both the top two PCs (Supp Figure 13a) and the T-SNE plot (Supp Figure 13b). Overall, the SAE-based model demonstrates less distinct separation between normal BMI controls and high-quality case pre-AOM periods (Supp Figure 14).

## Discussion

In this preliminary work, we outlined a workflow for obtaining EHR-based fine-grained obesity phenotypes at per patient visit level prior to initiating AOM therapy. Since EHR chronicled a patient’s physiological and pathological status, this may potentially help with AOM treatment decision making. This study aligns with the growing interest in classifying obesity into distinct subtypes and the adoption of the plural term “obesities” ^46,47^. It also lays the foundation for using EHR-based digital fingerprints as an alternative to traditional phenotyping approaches, which have shown potential in certain contexts ^27,48^ but not yet widely adopted by healthcare providers for precision obesity treatment due to their labor-intensive nature and insufficient granularity ^32^.

Our analysis identified at least nine distinct clusters before AOM initiation. Five clusters show clear clinical relevance independent of traditional obesity diagnoses (e.g., extreme obesity, localized adiposity). Below is a brief overview of the clinical significance of these clusters: **Cluster 1** primarily includes patients with chronic kidney disease (CKD) and metabolic disorders. The relationship between obesity and CKD is bidirectional ^49^: CKD can lead to water retention and increased body weight, while obesity promotes hyperfiltration and places a greater burden on the kidneys. These interactions are critical in selecting appropriate anti-obesity therapies ^50^. **Cluster 5** represents patients with obesity-related hyperlipidemia, characterized by elevated levels of atherogenic lipoproteins and higher blood pressure, but without severe comorbidities compared to other clusters. This cluster also shows the fewest observed clinical measurements, likely due to fewer comorbidities and less frequent clinical visits. **Cluster 6** is defined by reproductive and genital health disorders, which are often linked to hormonal imbalances and insulin resistance. The interaction between obesity and reproductive disorders, as well as the role of pharmacological interventions in weight loss, has been extensively studied ^51^. **Cluster 7** likely represents obese individuals with a higher susceptibility to obesity-related malignancies who were enrolled in weight management programs ^52^. Notably, the remarkably low frequency of cancer-related lab tests in this cluster suggests that these patients may have received cancer care at a different institution. The remarkably lower inflammatory response of **Cluster 9** may be caused by less visceral adiposity and more subcutaneous adiposity. This may be an overall healthier cluster with lower threshold for seeking treatment. These observations highlight the method’s potential to uncover unique patient groups, marking an important first step in digital phenotyping. Future steps would be to test cluster stability in a larger cohort, and incorporate additional environmental, psychological, behavioral, functional factors, and even microbiome profiles or genetic markers. Finally, linking these phenotypes to treatment responses would provide more comprehensive insights and reveal any subtle subgroup patterns.

The EHR-based clusters emphasize clinical manifestations and comorbidities, whereas traditional phenotyping, as described by ^27^, classifies obesity within a behavioral and metabolic framework (i.e., hungry brain, emotional hunger, hungry gut, and slow burn). While there is no exact mapping between the two, certain overlaps exist. For example, **Cluster 1** shares features with both the hungry brain and hungry gut phenotypes, reflecting advanced metabolic dysfunction (e.g., diabetes, coronary atherosclerosis) and CKD associated with chronic overeating. **Cluster 5** aligns with the emotional hunger phenotype, given its hyperlipidemia, high levels of atherogenic lipoproteins, elevated blood pressure, and fewer comorbidities. **Cluster 9** corresponds to the slow burn phenotype, with its low prevalence of malignancies, reduced systemic inflammation, preserved renal function, and absence of dramatic metabolic derangements, consistent with slower metabolic turnover. These potential overlaps suggest combining the strengths of each approach may provide a more complete understanding of obesity and its related disorders.

At this stage, we observed no obvious connection between visual separation in low-dimensional spaces, particularly the top two PCs, and clinical relevance. Even with advanced dimensionality reduction techniques T-SNE, there are generally no clear-cut boundaries between clinically distinct clusters. The findings highlight the challenges of visualizing complex diseases which often cannot be adequately captured in low-dimensional representations. Further research, such as MapperPlus ^53^ and other open frameworks ^54^, will be helpful to explore how to effectively visualize these clusters in a more clinically meaningful way beyond the static heatmap employed in current study.

We demonstrate the proposed GRU-D-AE architecture, along with its error function, effectively captures the nuances of high-dimensional, extremely sparse longitudinal EHR data. We also show that embeddings from native longitudinal data provide more clearly separated and distinct clusters than embeddings from static-transformed data. These characteristics make the architecture a highly valuable candidate for encoding longitudinal EHRs of obesity, as well as various other diseases. However, there are several caveats that deserve discussion and further investigation. First, the model exhibits a certain level of arbitrariness in handling missing values in different training folds. This behavior aligns with expectations, as the error function primarily minimizes the difference between input features (both observed and imputed) and reconstructed features. As a result, the model may exploit shortcuts (e.g. imputing all missing values as zero, as an extreme example) to satisfy the error function. A similar concern was raised by Daniel Jarrett et al. ^55^, who proposed target-embedding autoencoders, co-training the autoencoder with a target value to enhance stability. In another study ^56^, using target variables and clinical confounders improved the stability of embeddings in complex biomedical datasets. Nonetheless, selecting appropriate co-training targets (e.g., clinical outcomes, BMI trajectories, treatment responses) for deep phenotyping during pre-AOM periods remains challenging and is an important area for future investigation. Second, unlike the arbitrariness in missing data imputation, the top two PCs from the five training folds consistently formed similar clustering patterns, differing only in orientation. Moreover, examination of diagnosis and measurement characteristics of clusters in other training folds revealed highly consistent clinical traits (data not shown). These findings suggest that the bottleneck layer may capture essential temporal feature characteristics independent of the missing data imputation methods. Future studies should carefully assess the impact of imputation mechanisms on cluster stability.

In this study, we defined data quality as the observed proportion of lab or vital measurements within 1 year before AOM initiation. While this may be arbitrary given the anticipatable variation in feature availability across time and health care systems, it provides an intuitive reflection of how well a patient is documented by the EHR system during pre-AOM period. Given the observation that pre-AOM periods with above median measurement proportion (i.e 0.0453) formed clusters generally independent of data quality levels, we propose 5% as a cut-off threshold, below which the pre-AOM periods might be less distinguishable from each other. On the other hand, further research is needed to carefully assess the source of these low quality pre-AOM periods (e.g., limited access to healthcare, first-time visits, fewer comorbidities).

The laboratory and vital measurements used in this study are derived from the most commonly available features among obesity patients in the current EHR system. However, feature availability can vary over time and across healthcare systems. This variability, combined with the high dimensionality of EHR data, poses challenges for ensuring feature consistency, which can significantly affect the stability and reliability of clustering results. Bellman first introduced this issue in 1957, referring to it as the “curse of dimensionality” ^57^. Similar challenges ^58,59,60,61^ are prevalent in the medical field, where large feature sets and limited sample sizes are common. Specifically, Loftus et al. ^60^ reviewed these challenges in disease phenotyping, highlighting the importance of cohort representativeness and discussed caveats in data outliers, missing data, categorical variables, scaling, data transformation, and feature selection. For obesity, while certain strategies may improve cluster stability and potential clinical utility in the pre-AOM period, substantial challenges remain. Further research combining large cohorts with diverse demographic and SDOH backgrounds, and intricately designed clustering approaches, is needed to generate a holistic view that can inform clinical practice.

## Conclusion

Obesity is a complex, chronic condition, and its multifaceted nature poses significant challenges for deep phenotyping and precision medicine. Here we demonstrated that longitudinal EHR data can potentially serve as a valuable resource for deep phenotyping during the pre-AOM period at the individual patient visit level. Our analysis revealed the presence of clusters with distinct clinical significance, which could have an implication on AOM treatment options. Further research using larger, independent cohorts is necessary to validate the reproducibility and clinical relevance of these clusters, uncover more detailed substructures, and assess cluster-specific responses to AOM treatment.

## Limitations

In this exploratory study, we only shed light on a few limited aspects of obesity deep phenotyping from the perspective of EHR. Below, we outline key limitations and suggest areas for future research. First, although Houston is one of the most ethnically and racially diverse cities in the U.S., this study is still subject to the limitations inherent in a single-site study, such as selection bias, environmental and institutional influences, and regulatory differences. Broader studies conducted at the state or national level are necessary to validate the generalizability of the findings and to uncover more nuanced clustering structures. Second, due to the limited sample size, we only considered lab/vital measurements, and diagnosis codes for deep phenotyping, which does not provide comprehensive coverage of the factors that may influence obesity phenotypes (e.g., age, gender, race, ethnicity, medications, social determinants of health, and genetic/epigenetic information). Future research should include these factors as phenotyping features or examine clustering patterns across different subgroups (e.g., age subgroups). Third, this study focuses on clustering patterns during the pre-AOM period, but it does not explore how different clusters respond to various AOMs in terms of outcomes such as changes in body weight, BMI, lipid levels, and side effects. Given the numerous types, combinations, and exposure lengths of AOMs, this area needs to be thoroughly investigated in larger cohorts. Finally, we demonstrate that data quality significantly influences the visual separability of the top two PCs. However, due to space limitations, we did not delve deeply into the low-quality pre-AOM periods. Further investigation is needed to understand the underlying factors contributing to these data points (e.g., limited access to healthcare, first-time visits, fewer comorbidities) and their effect on the overall effectiveness of EHR-based deep phenotyping.

## Data availability

The data supporting this study are not publicly available due to restrictions related to patient privacy and confidentiality. Access to the de-identified UT-Physician data may be granted upon reasonable request and with appropriate institutional approvals.

## Contributions

XR led the study design, conducted data analysis, and drafted the manuscript. SL conducted data analysis on atemporal clustering. LW contributed to manuscript preparation. AW oversaw the harmonization of the UT-Physician EHR with the OMOP CDM. MS provided expert insights as an obesity clinician. HL contributed to the study design and coordinated the overall project.

## List of abbreviations

AOM: Anti-Obesity Medication
BMI: Body Mass Index
CCS: Clinical Classifications Software
CDM: Common Data Model
EHR: Electronic Health Record
F-AOM: FDA-approved AOM
FDA: Food and Drug Administration
GMM: Gaussian Mixture Model
HCP: Healthcare Practitioner
O-AOM: Off-label AOM
PCA: Principal Component Analysis
PC: Principal Component
UT: University of Texas

