## Supplementary Figures and Text for "Deep phenotyping obesity using EHR data: Promise, Challenges, and Future Directions"

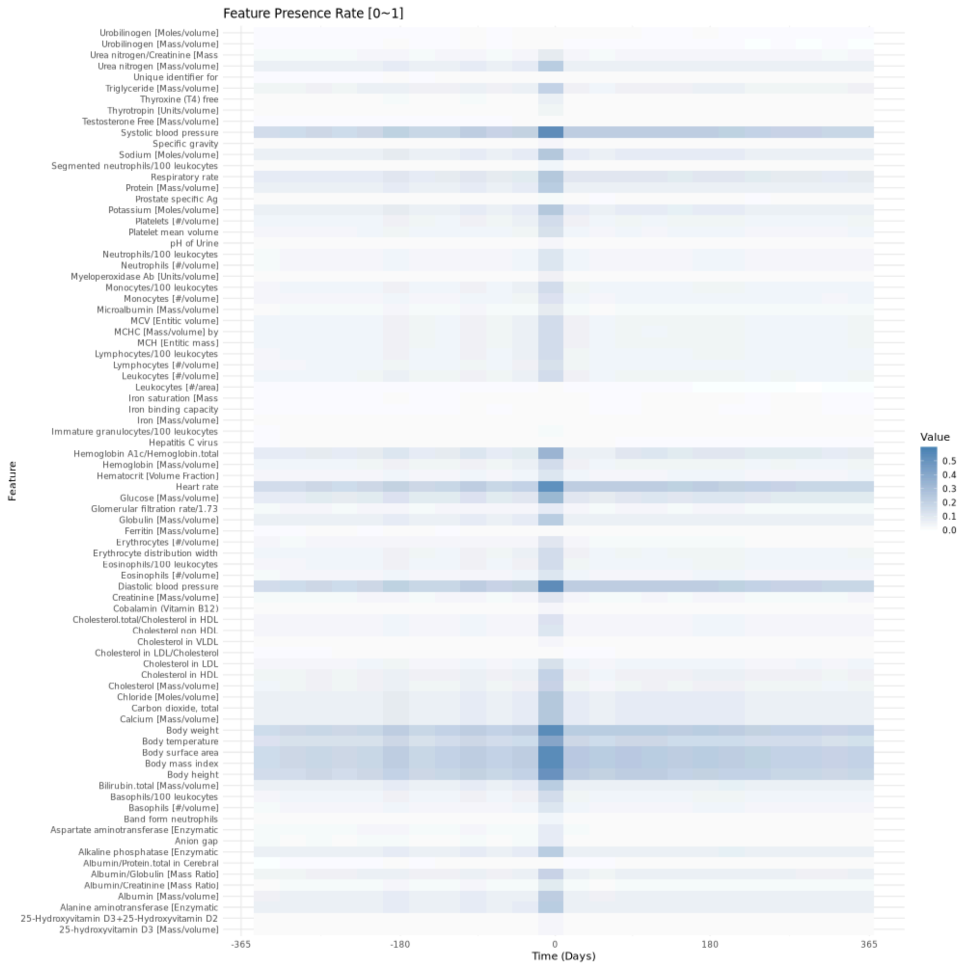


**Supp Figure 1** Measurement presence rate within 1 year before (i.e. pre-AOM period) and after initiating medium to long term AOM therapy.

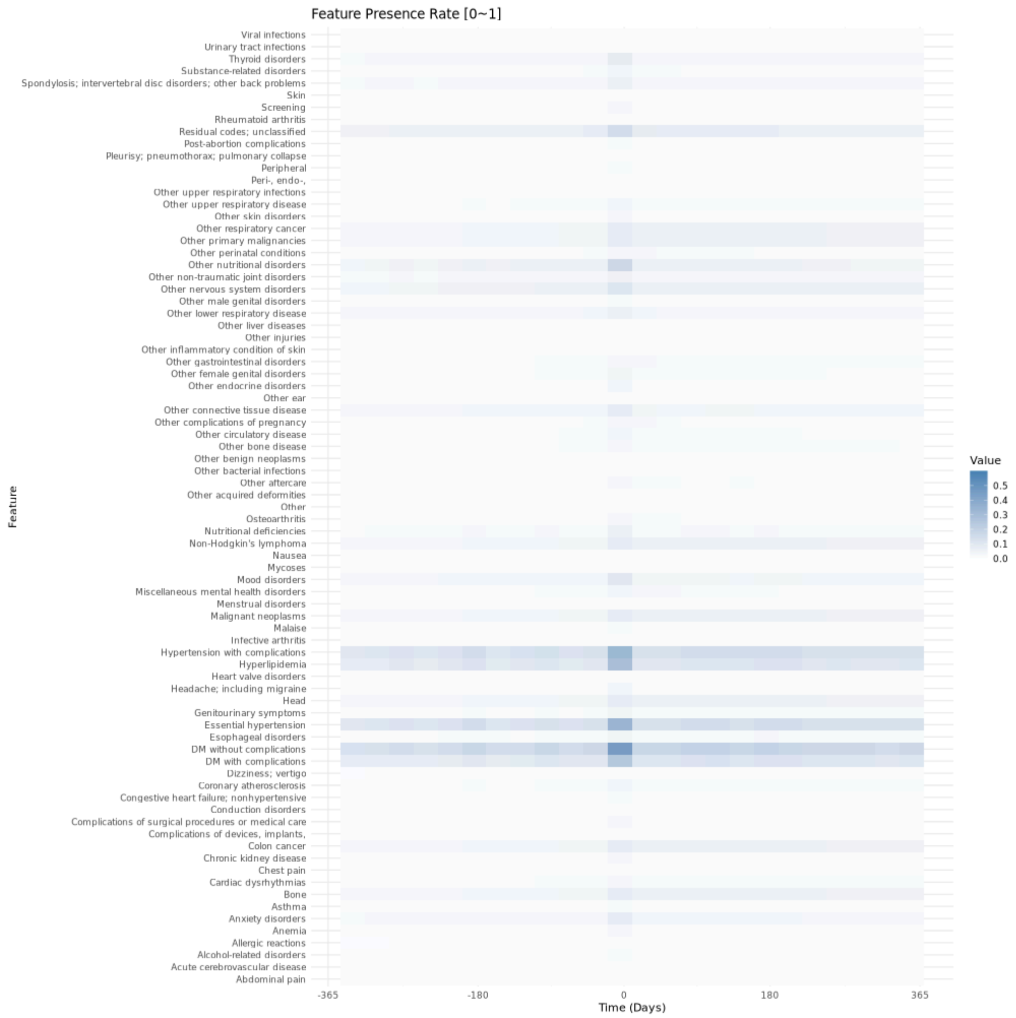


**Supp Figure 2** CCS code presence rate within 1 year before (i.e. pre-AOM period) and after initiating medium to long term AOM therapy.

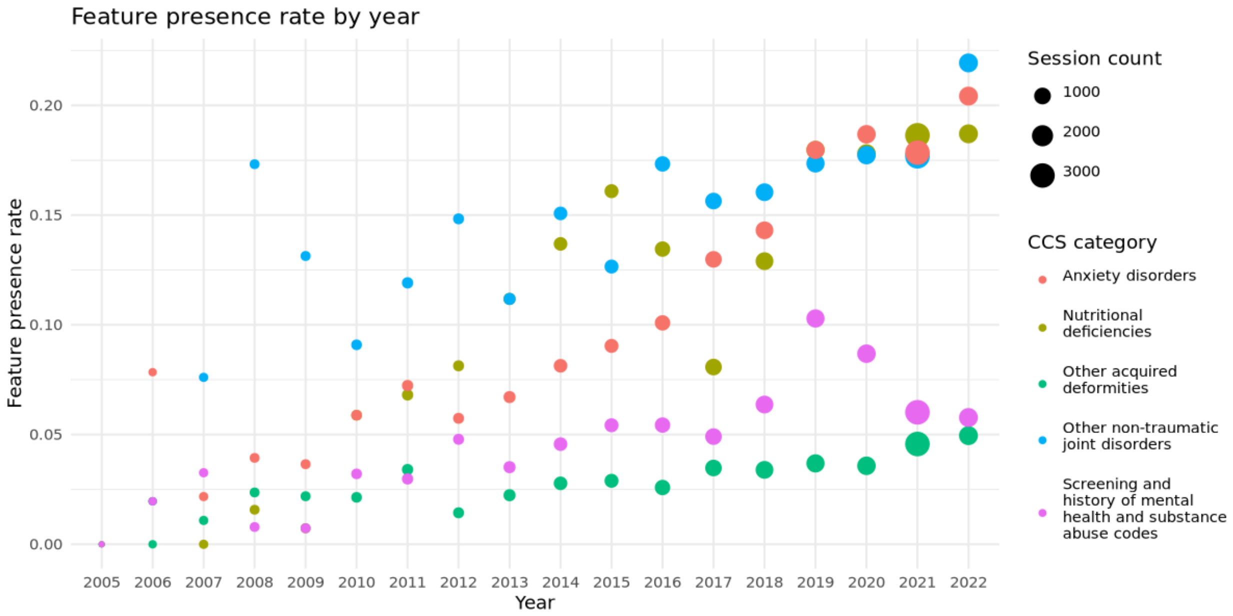


**Supp Figure 3** Top 5 CCS categories with significantly increased presence rate throughout years.

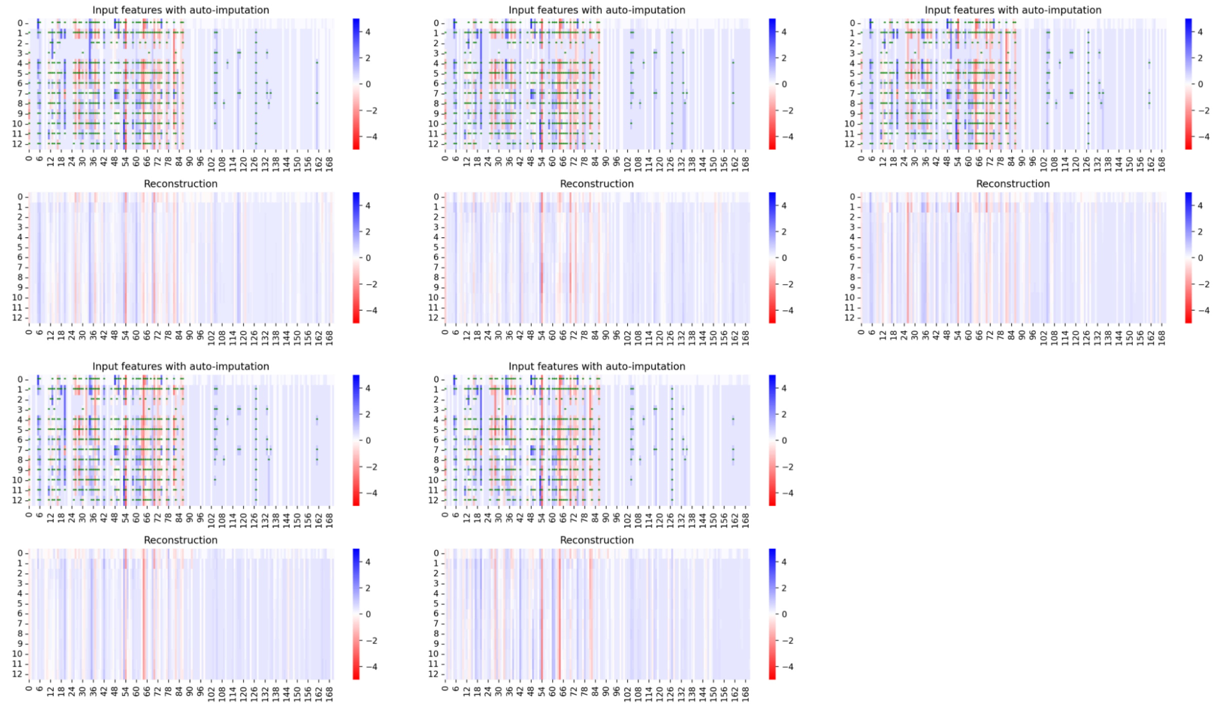


**Supp Figure 4** GRU-D-AE-based auto-imputation and reconstruction of a single patient's EHR profile during the pre-AOM period. The five subplots correspond to the outcomes of a 5-fold cross-validation using a leave-one-fold-out training approach. In each subplot, the upper section shows the original recorded values (green dots) and the imputed data for 171 longitudinal features (x-axis), each spanning 13 time points (y-axis) covering the period from 365 days to 5 days before AOM initiation. The lower section illustrates the reconstructed EHR profile after passing through a bottleneck layer consisting of 120 neurons.

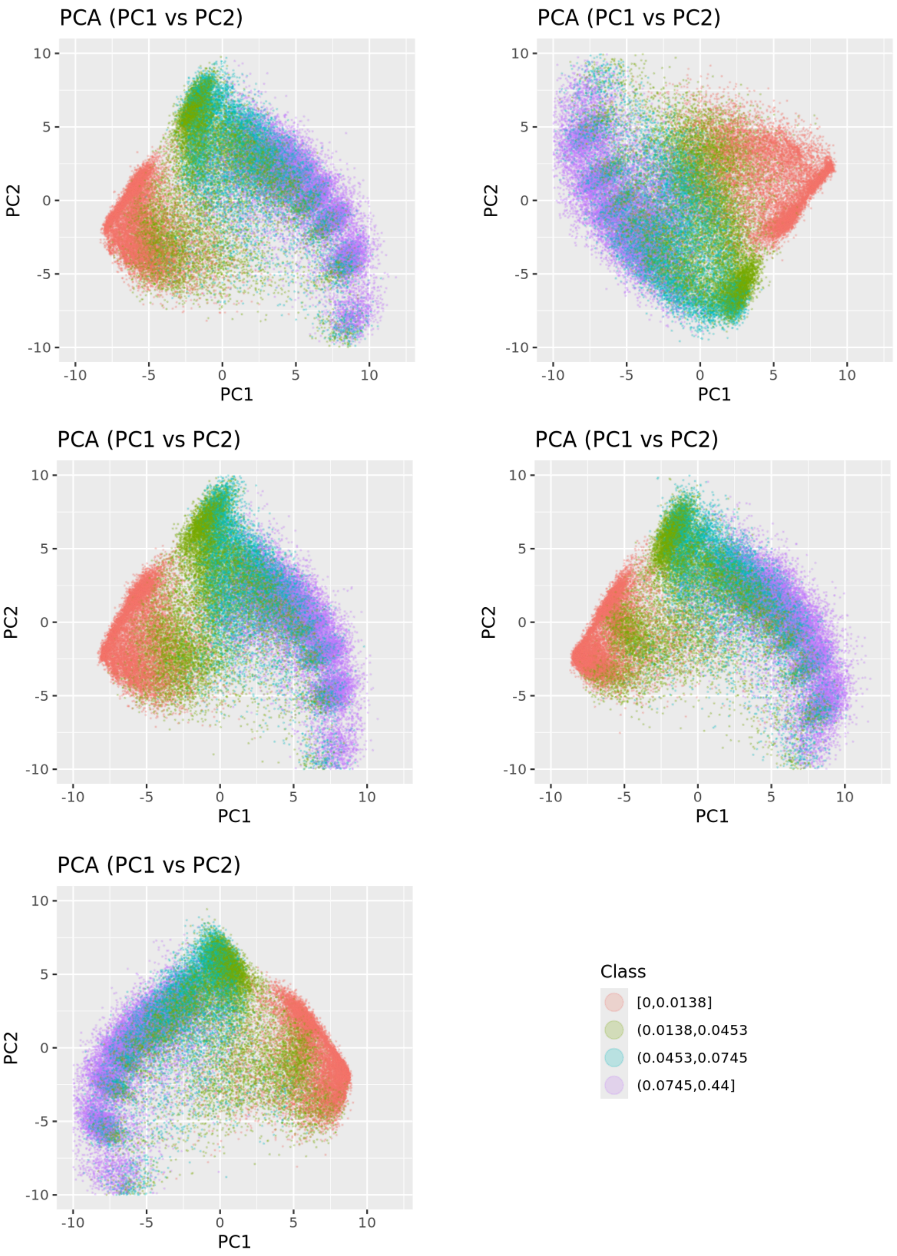


**Supp Figure 5** PCA-based clustering of all case embeddings from 5-fold models. Points colors were dimmed to highlight the cluster centers.

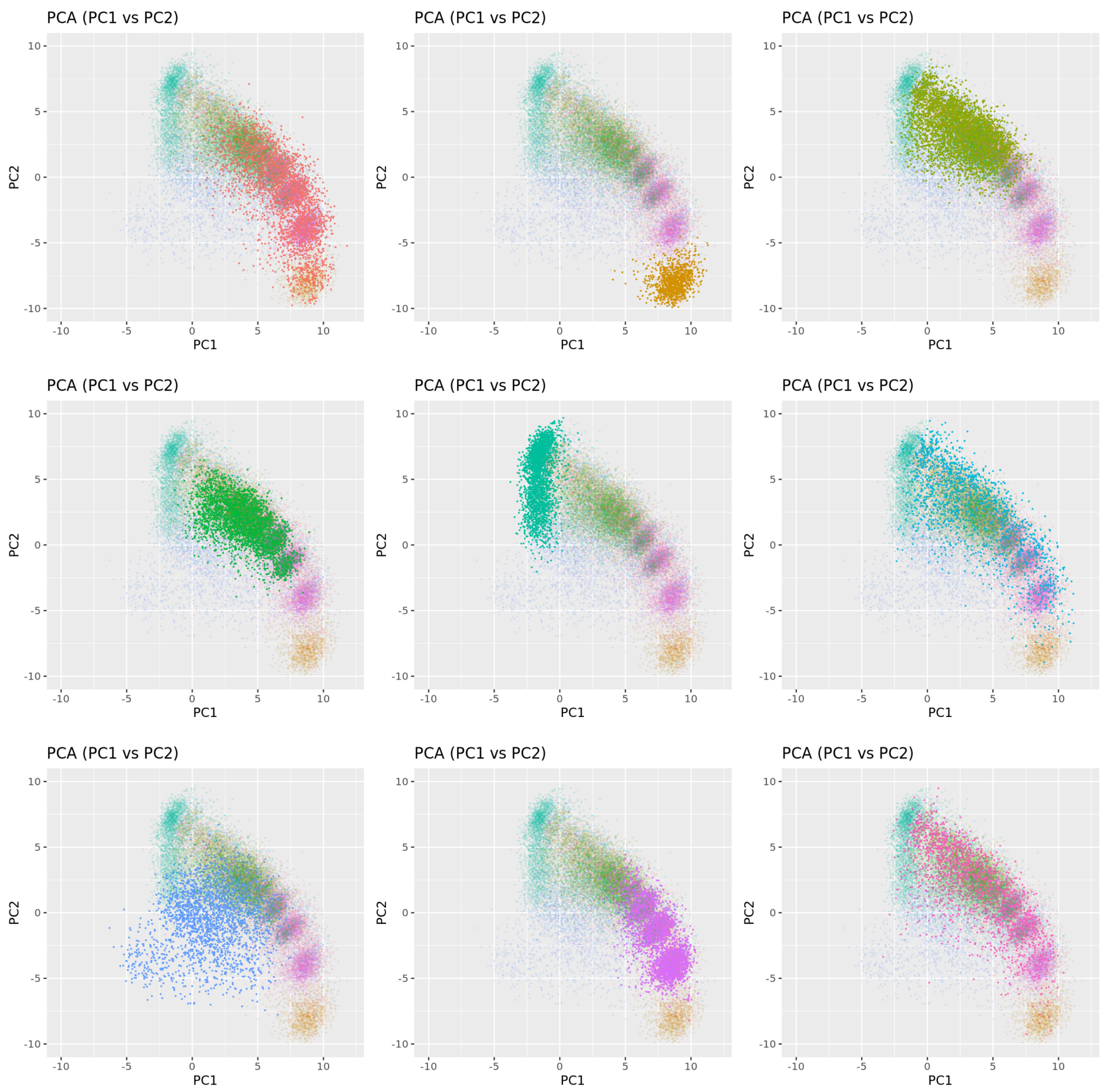


**Supp Figure 6** GMM identified nine clusters of high quality pre-AOM periods, highlighted in separate plots on the two major PCs' space.

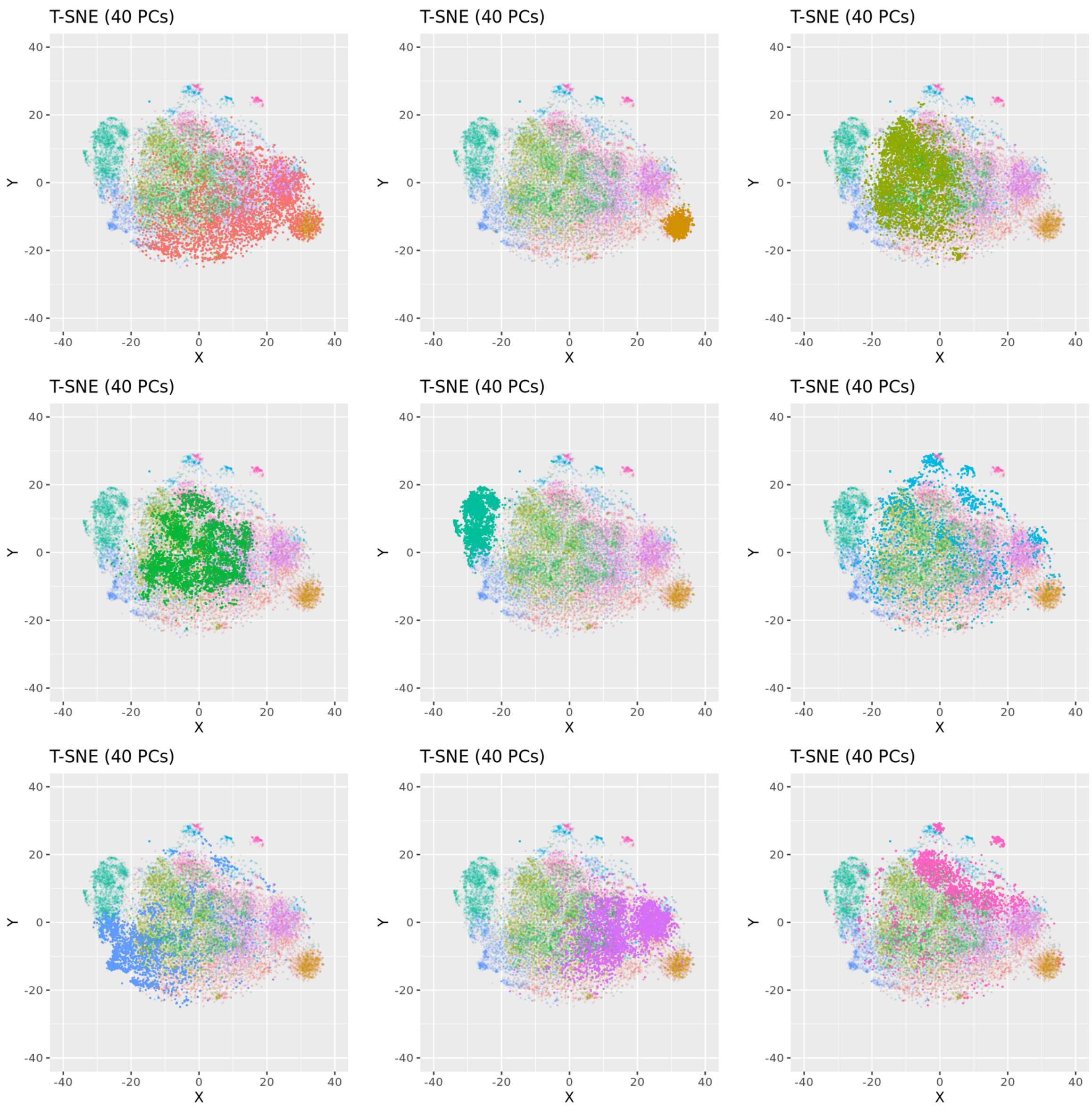


**Supp Figure 7** GMM identified nine clusters (filled by row) of high quality pre-AOM periods, highlighted in separate plots on T-SNE space.

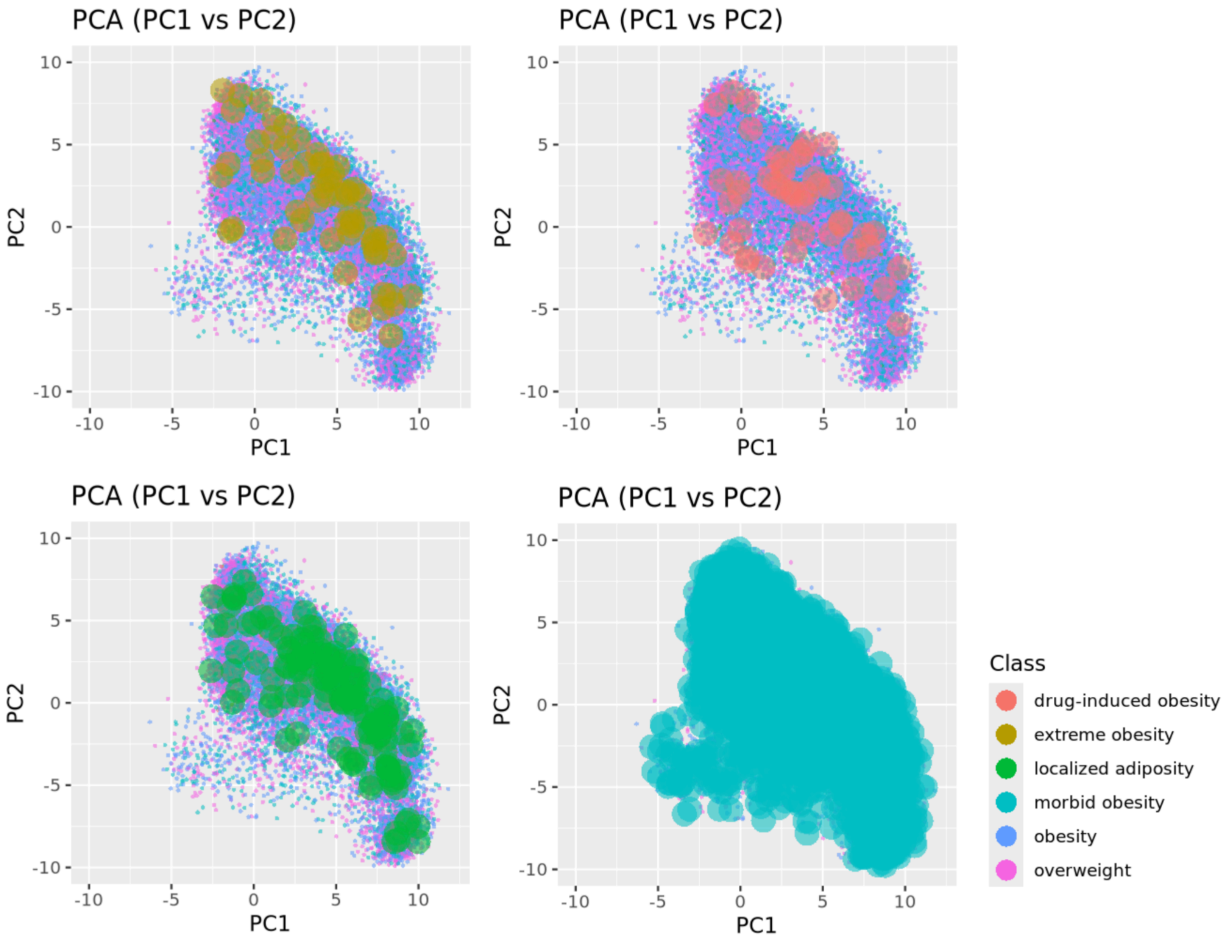


**Supp Figure 8** Distribution of traditional obesity phenotype in the context of GRU-D-AE-based clustering of pre-AOM periods.

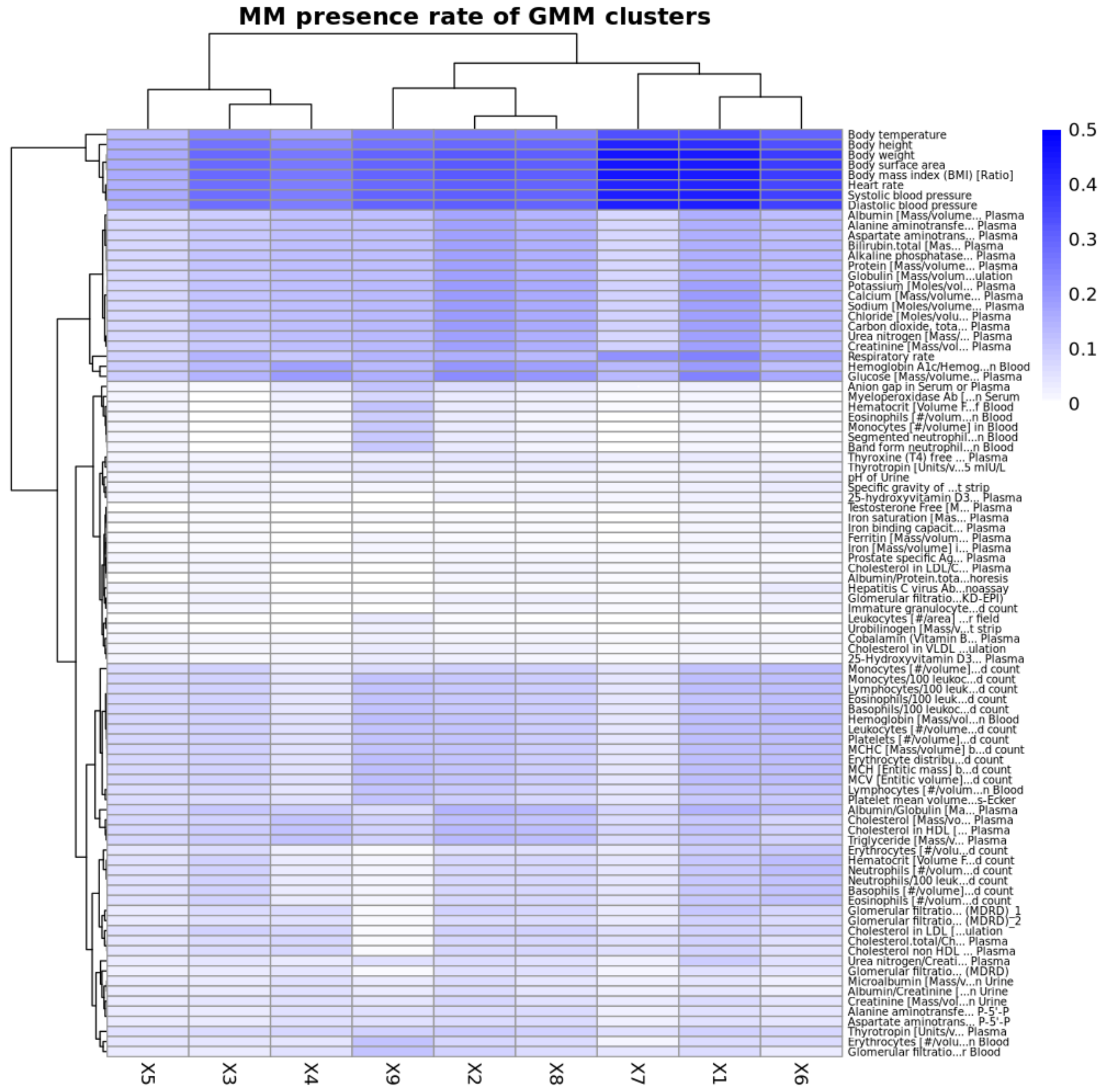


**Supp Figure 9** Average measurement presence rates against GMM-based clusters of pre-AOM periods. Presence rate is the proportion of measurements observed in the 13 time steps within 1 year before AOM initiation.

.

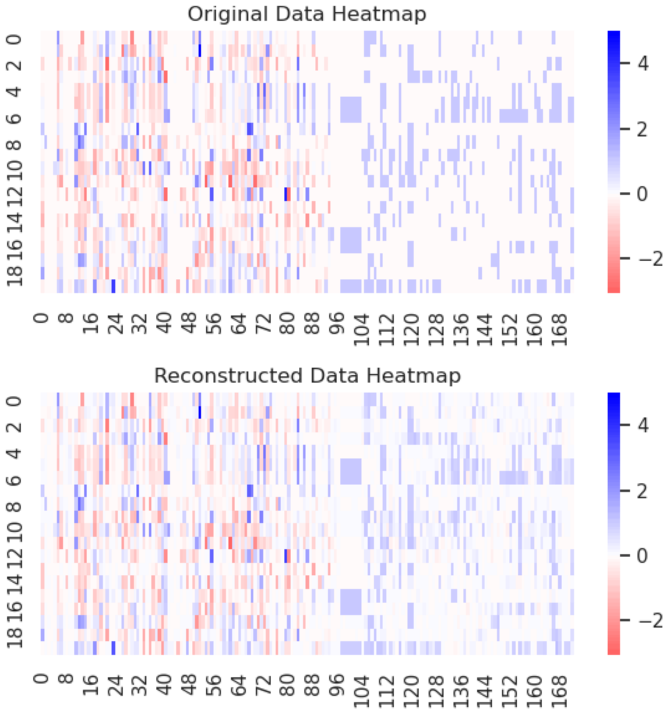


**Supp Figure 11** SAE-based reconstruction of the static transformed EHR profiles during the pre-AOM period for 20 patients (y-axis) across 171 features (x-axis). Result displayed for one of the 5-fold models, with other folds performing similarly.

a b

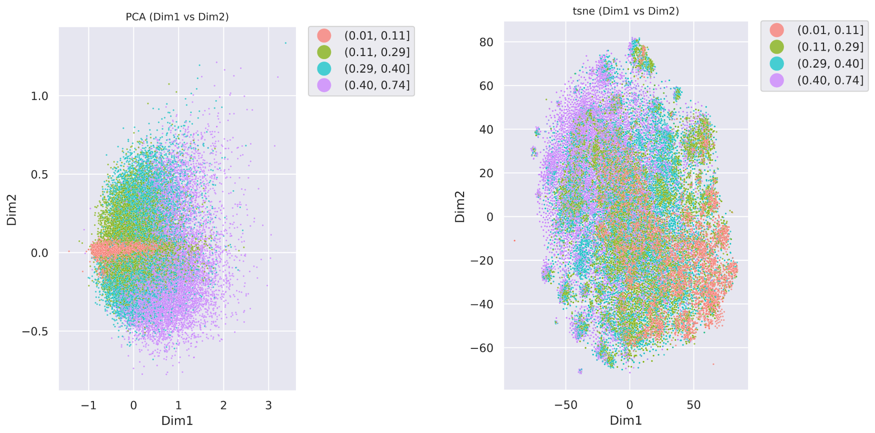


**Supp Figure 12** SAE-based clustering of case pre-AOM periods colored by data quality quartiles. a) Top two PCs, b) T-SNE plot. Result displayed for one of the 5-fold models, with other folds performing similarly.

a b

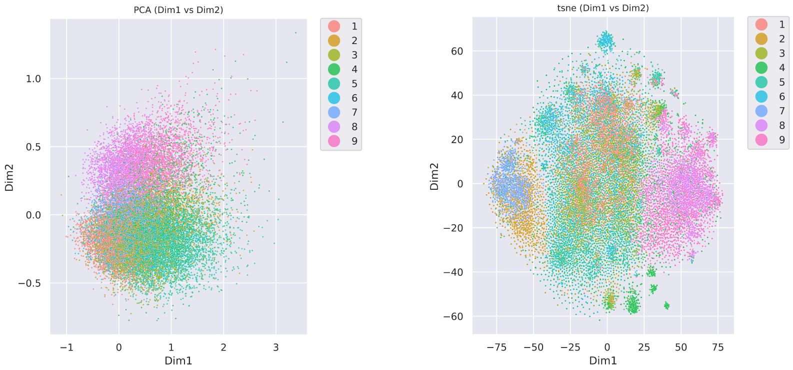


**Supp Figure 13** SAE-based clustering of high quality cases pre-AOM periods colored by GMM-based clusters. a) Top two PCs, b) T-SNE plot. Result displayed for one of the 5-fold models, with other folds performing similarly.

a b

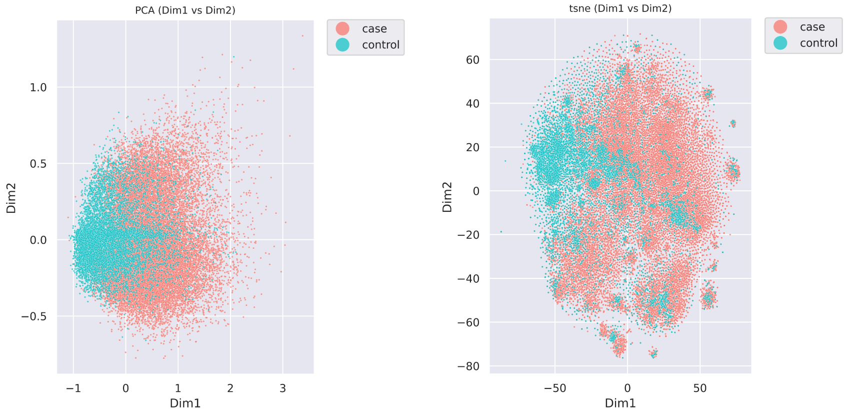


**Supp Figure 14** SAE-based clustering of normal BMI controls and high quality cases pre-AOM periods. a) Top two PCs, b) T-SNE plot. Result displayed for one of the 5-fold models, with other folds performing similarly.

**Supp Text 1 Concept codes for obesity diagnoses**

Obesity: 433736; Morbid obesity: 434005; Localized adiposity: 438731; Extreme obesity with alveolar hypoventilation: 4100857; Drug-induced obesity: 4097996; Simple obesity: 4217557

**Supp Text 2 Comorbidities for determining overweight**

prediabetes, diabetes, hypertension, metabolic syndrome, obstructive sleep apnea, polycystic ovary syndrome, insulin resistance, hyperlipidemia, fatty liver, non-alcoholic steatohepatitis, coronary artery disease, cerebrovascular accident, stroke, peripheral vascular disease, congestive heart failure, colon/breast/renal/endometrial/liver cancer, osteoarthritis, or with HbA1c >= 5.7 and < 6.5
